# Proportion of Hospitalizations Preventable with Increased Oral SARS-CoV-2 Antiviral Treatment

**DOI:** 10.1101/2023.12.19.23300241

**Authors:** Matthew E. Levy, Vanessa Chilunda, Phillip R. Heaton, Joseph Grzymski, Pamala A. Pawloski, Jason D. Goldman, Shishi Luo

## Abstract

We estimated the proportion of hospitalizations that could have been averted had all eligible high-risk adults with SARS-CoV-2 infection in a clinical cohort been treated with an oral SARS-CoV-2 antiviral agent early in infection. Among 3,037 patients with risk factors for progressing to severe COVID-19, 946 (31.1%) received an oral antiviral prescription (834 nirmatrelvir and 112 molnupiravir). Only 3.0% of treated patients vs 9.1% of untreated patients were hospitalized (adjusted risk ratio [RR]=0.27; 95% confidence interval [CI]: 0.18-0.41). If all patients had been treated, an estimated 63.3% (95% CI: 50.4-75.1) of hospitalizations within 30 days could have been prevented. This finding is attributed to large gaps in treatment and a strong protective association of antiviral treatment with subsequent hospitalization.

## Introduction

The oral SARS-CoV-2 antiviral agents nirmatrelvir (plus ritonavir) and molnupiravir are effective at preventing COVID-19-associated hospitalization.^1,2^ However, prescribing gaps exist among nonhospitalized SARS-CoV-2-infected patients with risk factors for progressing to severe COVID-19.^3^ To demonstrate the potential impact of improving guideline-concordant oral antiviral prescribing, we estimated the proportion of hospitalizations that could have been averted had all eligible individuals in a clinical cohort been treated early in infection.

## Methods

We performed a retrospective analysis of electronic health record data obtained from two health systems: HealthPartners (Minnesota and Wisconsin) and Renown Health (Nevada). This analysis included adults aged ≥18 years with a positive health system-administered SARS-CoV-2 test (molecular or antigen) between April 2022-June 2023 who had ≥1 outpatient visit in the preceding year and ≥1 risk factor for severe COVID-19 (≥50 years old, unvaccinated, or ≥1 high-risk condition).^4^ Detailed methods are provided in eMethods and eTables 1-3.

The index date was the collection date of each patient’s first positive test. All-cause hospitalization was defined as admission 0-30 days after the index date. Patients first tested on the date of hospitalization were included to minimize selection bias. Treatment was defined as receiving a prescription for nirmatrelvir or molnupiravir within 14 days before through 5 days after the index date (and, if hospitalized, ≥1 day pre-admission).

Patients with hospitalizations 1-30 days earlier or with outpatient remdesivir or SARS-CoV-2 monoclonal antibody treatment were excluded.

Modified Poisson regression was employed to estimate adjusted risk ratios (RRs) for the association between treatment and hospitalization, overall and within demographic and clinical subgroups.^5^ Applying the formula for population attributable fraction to the comparison of no treatment vs treatment (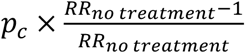, where *p* is the proportion of hospitalized cases who had not been treated), we estimated the ‘preventable fraction’—the proportion of hospitalizations that could be prevented if all persons were treated.^6^ Confidence intervals (CIs) were computed using the 2.5th and 97.5th percentiles from 1000 bootstrap resampling iterations. Analyses were performed using R v4.2.3.

## Results

Among 3,037 SARS-CoV-2-infected patients with risk factors for severe COVID-19, 35.7% were ≥65 years old, 20.9% were unvaccinated, 12.8% were immunocompromised, and 11.8% had Charlson comorbidity index (CCI) scores ≥3 (Table 1). Overall, 946 (31.1%) received oral antiviral prescriptions (834 nirmatrelvir; 112 molnupiravir).

**Table 1.**
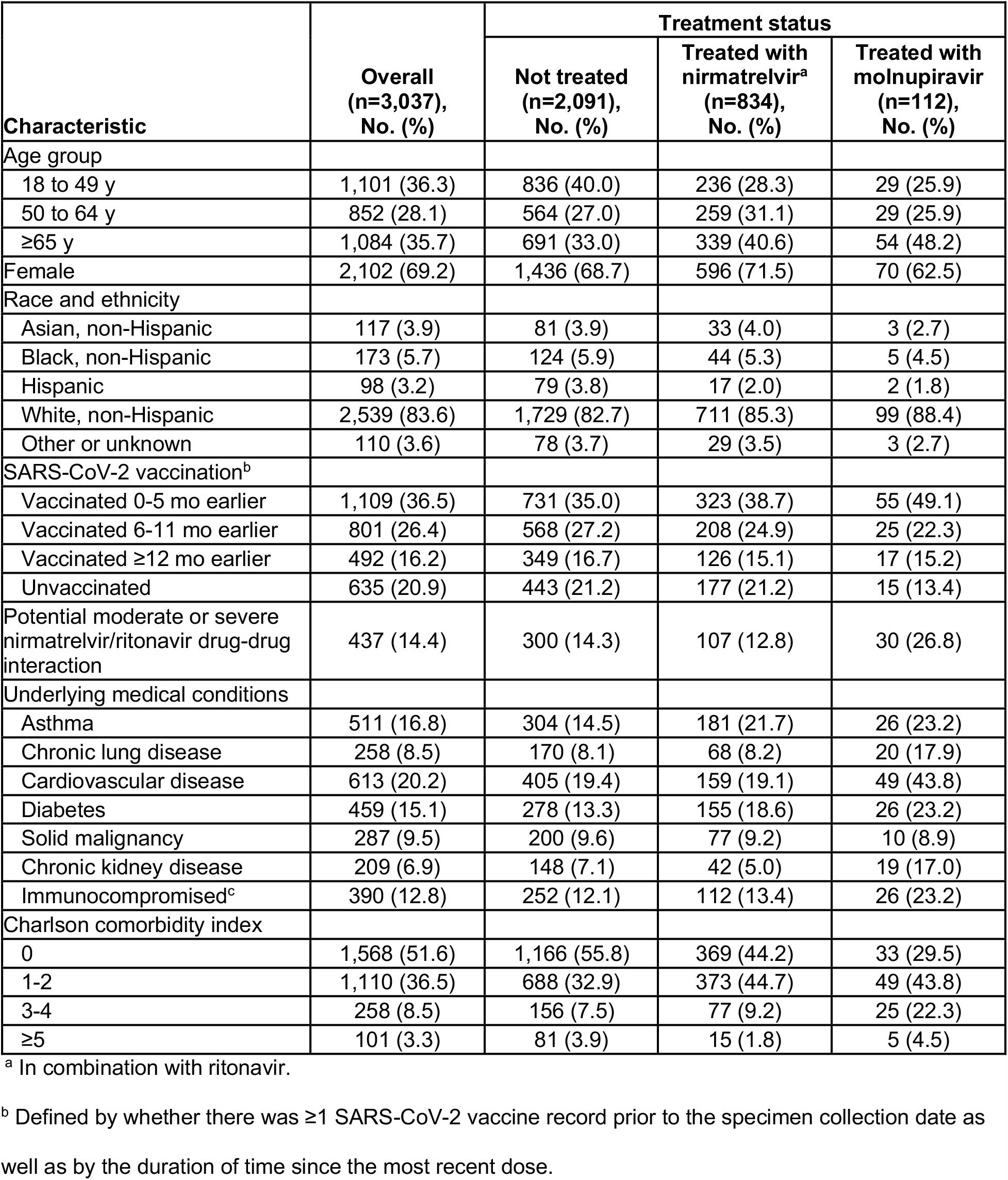
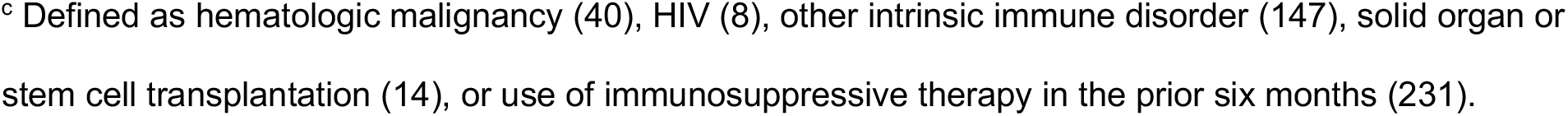
Patient Characteristics Overall and Stratified by Oral SARS-CoV-2 Antiviral Treatment Status.

| Characteristic | Overall<br>(n=3,037),<br>No. (%) | Treatment status |  |  |
| --- | --- | --- | --- | --- |
|  |  | Not treated<br>(n=2,091),<br>No. (%) | Treated with<br>nirmatrelvir<sup>a</sup><br>(n=834),<br>No. (%) | Treated with<br>molnupiravir<br>(n=112),<br>No. (%) |
| Age group |  |  |  |  |
| 18 to 49 y | 1,101 (36.3) | 836 (40.0) | 236 (28.3) | 29 (25.9) |
| 50 to 64 y | 852 (28.1) | 564 (27.0) | 259 (31.1) | 29 (25.9) |
| ≥65 y | 1,084 (35.7) | 691 (33.0) | 339 (40.6) | 54 (48.2) |
| Female | 2,102 (69.2) | 1,436 (68.7) | 596 (71.5) | 70 (62.5) |
| Race and ethnicity |  |  |  |  |
| Asian, non-Hispanic | 117 (3.9) | 81 (3.9) | 33 (4.0) | 3 (2.7) |
| Black, non-Hispanic | 173 (5.7) | 124 (5.9) | 44 (5.3) | 5 (4.5) |
| Hispanic | 98 (3.2) | 79 (3.8) | 17 (2.0) | 2 (1.8) |
| White, non-Hispanic | 2,539 (83.6) | 1,729 (82.7) | 711 (85.3) | 99 (88.4) |
| Other or unknown | 110 (3.6) | 78 (3.7) | 29 (3.5) | 3 (2.7) |
| SARS-CoV-2 vaccination<sup>b</sup> |  |  |  |  |
| Vaccinated 0-5 mo earlier | 1,109 (36.5) | 731 (35.0) | 323 (38.7) | 55 (49.1) |
| Vaccinated 6-11 mo earlier | 801 (26.4) | 568 (27.2) | 208 (24.9) | 25 (22.3) |
| Vaccinated ≥12 mo earlier | 492 (16.2) | 349 (16.7) | 126 (15.1) | 17 (15.2) |
| Unvaccinated | 635 (20.9) | 443 (21.2) | 177 (21.2) | 15 (13.4) |
| Potential moderate or severe<br>nirmatrelvir/ritonavir drug-drug<br>interaction | 437 (14.4) | 300 (14.3) | 107 (12.8) | 30 (26.8) |
| Underlying medical conditions |  |  |  |  |
| Asthma | 511 (16.8) | 304 (14.5) | 181 (21.7) | 26 (23.2) |
| Chronic lung disease | 258 (8.5) | 170 (8.1) | 68 (8.2) | 20 (17.9) |
| Cardiovascular disease | 613 (20.2) | 405 (19.4) | 159 (19.1) | 49 (43.8) |
| Diabetes | 459 (15.1) | 278 (13.3) | 155 (18.6) | 26 (23.2) |
| Solid malignancy | 287 (9.5) | 200 (9.6) | 77 (9.2) | 10 (8.9) |
| Chronic kidney disease | 209 (6.9) | 148 (7.1) | 42 (5.0) | 19 (17.0) |
| Immunocompromised<sup>c</sup> | 390 (12.8) | 252 (12.1) | 112 (13.4) | 26 (23.2) |
| Charlson comorbidity index |  |  |  |  |
| 0 | 1,568 (51.6) | 1,166 (55.8) | 369 (44.2) | 33 (29.5) |
| 1-2 | 1,110 (36.5) | 688 (32.9) | 373 (44.7) | 49 (43.8) |
| 3-4 | 258 (8.5) | 156 (7.5) | 77 (9.2) | 25 (22.3) |
| ≥5 | 101 (3.3) | 81 (3.9) | 15 (1.8) | 5 (4.5) |

<sup>a</sup> In combination with ritonavir.
<sup>b</sup> Defined by whether there was ≥1 SARS-CoV-2 vaccine record prior to the specimen collection date as well as by the duration of time since the most recent dose.
<sup>c</sup> Defined as hematologic malignancy (40), HIV (8), other intrinsic immune disorder (147), solid organ or stem cell transplantation (14), or use of immunosuppressive therapy in the prior six months (231).

Only 3.0% of treated patients vs 9.1% of untreated patients were hospitalized (Table 2). Among 218 hospitalizations, 163 (74.8%) occurred on the date of SARS-CoV-2 testing. After adjustment, treatment was associated with reduced risk of hospitalization for any oral antiviral agent (RR=0.27; 95% CI: 0.18-0.41), nirmatrelvir (RR=0.36; CI: 0.22-0.58), and molnupiravir (RR=0.22; CI: 0.08-0.59). The association for any treatment was stronger in patients aged ≥65 (RR=0.15) vs 18-49 (RR=0.48) years (interaction, p<0.001).

**Table 2.**
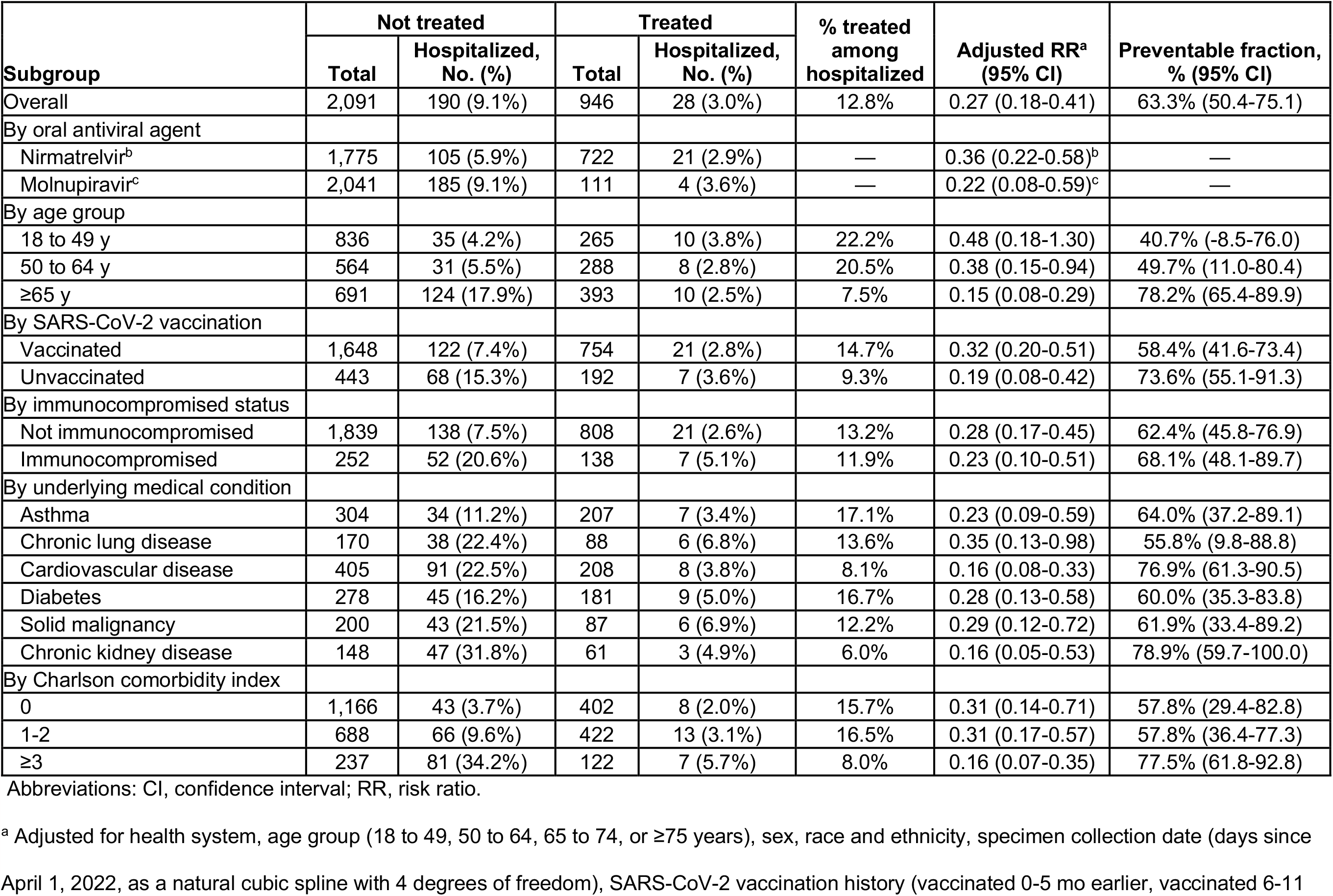
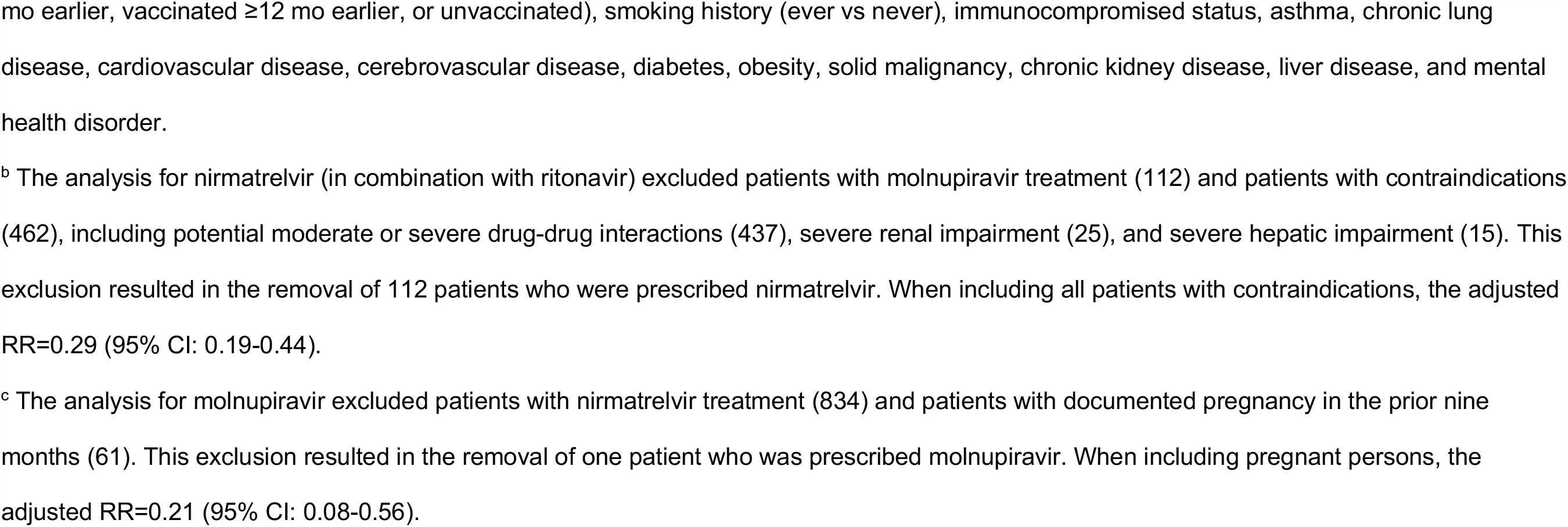
Association of Oral SARS-CoV-2 Antiviral Treatment with Hospitalization and the Preventable Fraction.

If all patients had been treated, an estimated 63.3% (95% CI: 50.4-75.1) of hospitalizations could have been averted (Table 2). The preventable fraction was similarly high in subgroups who were older (≥65 years) (78.2%; CI: 64.8-89.9), unvaccinated (73.6%; CI: 55.1-91.3), immunocompromised (68.1%; CI: 48.1-89.7), or with multimorbidity (CCI ≥3) (77.5%; CI: 61.8-92.8).

## Discussion

Among patients testing positive for SARS-CoV-2 with high risk for COVID-19 progression, the majority of hospitalizations within 30 days could have been prevented if all patients had received early antiviral treatment. This finding is attributed to large gaps in treatment and a strong protective association of antiviral treatment with subsequent hospitalization. Given that many hospitalizations occurred on the date of testing, underutilization of outpatient testing or limited access to treatment upon onset of symptoms may be an additional contributing factor to underuse of antivirals. Study limitations include the inability to confirm symptomatic COVID-19, lack of data on timing of symptom onset and causes of hospitalization, and reliance on prescriptions for defining antiviral treatment.

## Data Availability

Data produced in the present study are not available due to data sharing agreements with partner institutions.

## Additional Contributions

We acknowledge the Helix Clinical Informatics and Bioinformatics teams for their contributions to electronic health record data and viral sequencing pipelines. We also acknowledge Catherine Clinton for her oversight and guidance in ensuring research compliance. We thank HealthPartners, Renown Health, and the Desert Research Institute for helping to launch the Helix Cohorts, and we thank all collaborators and research participants within the ViEW Network™, Healthy Nevada Project, and myGenetics protocols.

## References

1. Lin DY, Abi Fadel F, Huang S, et al. Nirmatrelvir or molnupiravir use and severe outcomes from omicron infections. JAMA Netw Open. 2023;6(9):e2335077. doi:10.1001/jamanetworkopen.2023.35077

2. Paraskevis D, Gkova M, Mellou K, et al. Real-world effectiveness of molnupiravir and nirmatrelvir/ritonavir as treatments for COVID-19 in high-risk patients. J Infect Dis. Published online August 11, 2023. doi:10.1093/infdis/jiad324

3. Yan L, Streja E, Li Y, et al. Anti–SARS-CoV-2 Pharmacotherapies among nonhospitalized US veterans, January 2022 to January 2023. JAMA Netw Open. 2023;6(8):e2331249. doi:10.1001/jamanetworkopen.2023.31249

4. Underlying medical conditions associated with higher risk for severe COVID-19: information for healthcare professionals. Centers for Disease Control and Prevention. Updated February 9, 2023. Accessed November 27, 2023. https://www.cdc.gov/coronavirus/2019-ncov/hcp/clinical-care/underlyingconditions.html

5. Zou G. A modified poisson regression approach to prospective studies with binary data. Am J Epidemiol. 2004;159(7):702–706. doi:10.1093/aje/kwh090

6. Khosravi A, Nazemipour M, Shinozaki T, Mansournia MA. Population attributable fraction in textbooks: time to revise. Global Epidemiol. 2021;3:100062. doi:10.1016/j.gloepi.2021.100062

