## Supplemental Methods for "Proportion of Hospitalizations Preventable with Increased Oral SARS-CoV-2 Antiviral Treatment"

### Supplemental Online Content

**eMethods.**

**eTable 1.** Description of Electronic Health Record Data Sources.

**eTable 2.** Definitions of High-Risk Conditions.

**eTable 3.** Definitions of Nirmatrelvir/Ritonavir Contraindications.

**eMethods.**

**Data Source**

Electronic health record data were obtained from three study protocols which included two health systems with clinics spanning Minnesota and western Wisconsin (HealthPartners) and northern Nevada (Renown Health) (eTable 1). Data were collected using the Observational Medical Outcomes Partnership (OMOP) Common Data Model.[1] Study protocols were approved by institutional review boards (local or central) prior to study conduct.

**Analytic Sample**

This analysis included patients aged ≥18 years who had a positive molecular or antigen SARS-CoV-2 test result based on testing administered by the health system. Within each study protocol, each eligible patient could be included only once. If a patient had two or more positive SARS-CoV-2 test results during the study period, only the earliest test was used. Patients with an earlier positive SARS-CoV-2 test result within the prior 30 days – and prior to the start of the study period – were excluded (only relevant to patients with a positive test during the first 29 days of the study period for each protocol). Each patient's index date was defined as the date of specimen collection associated with their first SARS-CoV-2 test result during the study period.

The analytic sample was restricted to patients who had ≥1 risk factor for severe COVID-19 (88.2% of adult patients with SARS-CoV-2 infection), which included patients aged ≥50 years, patients with no known prior SARS-CoV-2 vaccination, and patients with ≥1 high-risk condition (as of their index date).[2] High-risk conditions included asthma, cardiovascular disease, cerebrovascular disease, chronic kidney disease, chronic lung disease, dementia, developmental or intellectual disability, diabetes, hematologic malignancy, HIV, liver disease, mental health condition, obesity, other intrinsic immune disorder, solid malignancy, solid organ or stem cell transplantation, smoking (current or former), or immunosuppressive medication use in the prior six months. Definitions for high-risk conditions are detailed in eTable 2.

To maximize the likelihood that patients would have accessed outpatient testing or treatment from the participating health system (as opposed to accessing outpatient medical care elsewhere and only engaging with the participating health system for severe COVID-19 requiring hospitalization), we further restricted the analytic sample to patients who had ≥1 outpatient visit in the year before their index date (89.9% of the patients with ≥1 risk factor for severe COVID-19). Outpatient visits included visits coded as any one of the following types: office visit, outpatient visit, outpatient hospital, ambulatory surgical center, urgent care facility, health examination, or observation room.

We further excluded patients who were hospitalized at any time during the 1-30 days before the index date to ensure that only new hospitalizations were included and that SARS-CoV-2 infection preceded hospitalization (115 [3.6%] excluded). Patients with outpatient prescriptions for both nirmatrelvir and molnupiravir (2) or with outpatient prescription/administration records for either remdesivir (5), a SARS-CoV-2 monoclonal antibody (bebtelovimab, sotrovimab, tixagevimab-cilgavimab, or bamlanivimab) (6), or COVID-19 convalescent plasma (0) were also excluded (13 [0.4%] excluded). In addition, seven patients with outpatient prescriptions for nirmatrelvir or molnupiravir 15-30 days before the index date (0.2%) and one patient with an outpatient prescription 6-30 days after the index date (<0.1%) were excluded. One patient who died within 30 days of the index date (without hospitalization) was also excluded (<0.1%).

**Outcome**

The outcome was new all-cause hospitalization occurring either on the index date or within 30 days after the index date. Although individuals with the outcome at time zero are typically excluded from cohort studies evaluating the risk of an incident outcome, we included patients admitted to the hospital on the same day as SARS-CoV-2 testing to minimize selection bias. The true time zero of interest is the initial date of symptom onset following SARS-CoV-2 infection, which is unknown in this study. However, the date of symptom onset is generally expected to be before the time of health system-administered testing, especially for patients whose first interaction with the health system while infected was hospital admission. Among all eligible hospitalizations, 74.8% occurred on the same day of testing (i.e., SARS-CoV-2 infection was likely first diagnosed upon being hospitalized). Thus, excluding these cases would exclude the majority of hospitalizations, which – in addition to making results less generalizable to real-world settings – may, on average, exclude the more severe cases that progressed more rapidly and/or the patients that were least likely to access outpatient testing or care earlier in the course of infection. The duration of time between symptom onset and testing is likely different between hospitalized and nonhospitalized patients, thus creating an imbalance when evaluating the timing of data points with respect to the index date. The impact of this methodological issue was minimized by including patients who were first tested for SARS-CoV-2 on the date of hospital admission as well as by restricting the sample to patients with outpatient visit(s) in the prior year and using a relatively wide window of 30 days for evaluating new hospitalizations (extending beyond the period of acute infection irrespective of the date of symptom onset). For the aforementioned reasons, hospitalization was defined as a dichotomous outcome rather than a time-to-event outcome.

**Oral SARS-CoV-2 Antiviral Treatment**

Treatment with an oral SARS-CoV-2 antiviral agent was defined as receipt of a prescription for nirmatrelvir (in combination with ritonavir) or molnupiravir within 14 days before through 5 days after the index date—and, if hospitalized 0-5 days after the index date, also at least one day before hospital admission. Similar to the rationale provided above regarding the timing of hospitalizations (see *Outcome*), the timing of antiviral prescriptions with respect to the testing date is less meaningful than the timing with respect to symptom onset (which is unknown) and the timing with respect to hospitalization (if hospitalized). We used a window extending back 14 days because recent prior treatment within that period is likely to have been related to acute infection associated with the index positive test result. We used a window extending forward five days to reflect the recommendation that nirmatrelvir or molnupiravir be taken within five days of onset of symptoms. Among the treated patients included, most received their prescription on either the day of the positive test (269; 28.4%) or one day after the positive test (536; 56.7%), whereas 83 (8.8%) received a prescription 2-5 days after the positive test, 52 (5.5%) received a prescription 1-5 days before the positive test, and 6 (0.6%) received a prescription 6-14 days before the positive test. Information on prescription fills was not available.

**Statistical Analysis**

Multivariable modified Poisson regression with robust error variance was employed to estimate adjusted risk ratios (RRs) and 95% confidence intervals (CIs) for the association between any oral SARS-CoV-2 antiviral treatment and subsequent incident all-cause hospitalization.[3] A model was created in the overall sample and within subgroups defined by age group, SARS-CoV-2 vaccination status, immunocompromised status, specific underlying medical conditions, and Charlson comorbidity index score.

The Charlson comorbidity index is a widely used algorithm for measuring the burden of disease from comorbidities that has predicted 1-year mortality risk.[4] It evaluates the presence of codes across 17 conditions: myocardial infarction, congestive heart failure, peripheral vascular disease, cerebrovascular disease, dementia, chronic pulmonary disease, rheumatologic disease, peptic ulcer disease, mild liver disease, diabetes without chronic complication, diabetes with chronic complication, hemiplegia or paraplegia, renal disease, any malignancy including lymphoma and leukemia, moderate or severe liver disease, metastatic solid tumor, or HIV. We implemented an adaptation of the Charlson comorbidity index calculation designed for *International Classification of Diseases, Tenth Revision, Clinical Modification* (ICD-10-CM) codes, as described by Beyrer et al.[5]

We also calculated separate adjusted RRs for nirmatrelvir treatment and molnupiravir treatment. For nirmatrelvir, we excluded patients treated with molnupiravir and patients with a nirmatrelvir/ritonavir contraindication, which included potential moderate or severe drug-drug interactions,[6] severe renal impairment, and severe hepatic impairment (eTable 3). For molnupiravir, we excluded patients treated with nirmatrelvir and patients with a recorded diagnosis, observation, procedure, or drug code representing a pregnancy, either on the index date or within the prior nine months. Pregnancy was evaluated among women aged 18-55 years and, within the OMOP Common Data Model, was defined as the documentation of a code that is either a descendant of or mapped to the SNOMED code 118185001, which represents 'Finding related to pregnancy'.

Models were adjusted for health system, age group (18 to 49, 50 to 64, 65 to 74, or ≥75 years), sex, race and ethnicity, specimen collection date (days since April 1, 2022, as a natural cubic spline with 4 degrees of freedom), SARS-CoV-2 vaccination history (vaccinated 0-5 months earlier, vaccinated 6-11 months earlier, vaccinated ≥12 months earlier, or unvaccinated), smoking history (ever vs never), immunocompromised status, asthma, chronic lung disease, cardiovascular disease, cerebrovascular disease, diabetes, obesity, solid malignancy, chronic kidney disease, liver disease, and mental health disorder.

Additionally, by reverse-coding treatment status (i.e., no treatment vs treatment) to obtain adjusted RRs for the comparison of no treatment vs treatment, and applying Miettenen's formula for population attributable fraction: $p_c \times \frac{adjusted\ RR_{no\ treatment} - 1}{adjusted\ RR_{no\ treatment}}$, where $p_c$ is the proportion of hospitalized cases who had not been treated, we estimated the 'preventable fraction' for any oral SARS-CoV-2 treatment—the proportion of all hospitalizations that would be prevented if the entire study population were treated with an oral SARS-CoV-2 antiviral agent.[7-9] The value for $p_c$ is equal to 1 minus the proportion of hospitalized patients who had been treated, and the adjusted RR for no treatment is equal to 1 divided by the adjusted RR for treatment. This epidemiologic measure relies on the assumption that the association between treatment and hospitalization is causal and that confounding is eliminated through adjustment. CIs for preventable fractions were computed using the 2.5th and 97.5th percentiles from 1000 bootstrap resampling iterations (with replacement).[10] Analyses were performed using R version 4.2.3.

**References**


1. Voss EA, Makadia R, Matcho A, et al. Feasibility and utility of applications of the common data model to multiple, disparate observational health databases. *J Am Med Inform Assoc*. 2015;22(3):553-564. doi:10.1093/jamia/ocu023
2. Underlying medical conditions associated with higher risk for severe COVID-19: information for healthcare professionals. Centers for Disease Control and Prevention. Updated February 9, 2023. Accessed November 27, 2023. https://www.cdc.gov/coronavirus/2019-ncov/hcp/clinical-care/underlyingconditions.html
3. Zou G. A modified poisson regression approach to prospective studies with binary data. *Am J Epidemiol*. 2004;159(7):702-706. doi:10.1093/aje/kwh090
4. Charlson ME, Pompei P, Ales KL, MacKenzie CR. A new method of classifying prognostic comorbidity in longitudinal studies: development and validation. *J Chronic Dis*. 1987;40(5):373-383. doi:10.1016/0021-9681(87)90171-8
5. Beyrer J, Manjelievskaia J, Bonafede M, et al. Validation of an International Classification of Disease, 10th revision coding adaptation for the Charlson Comorbidity Index in United States healthcare claims data. *Pharmacoepidemiol Drug Saf*. 2021;30(5):582-593. doi:10.1002/pds.5204
6. Drug-drug interactions between ritonavir-boosted nirmatrelvir (Paxlovid) and concomitant medications. National Institutes of Health. Updated November 2, 2023. Accessed November 28, 2023. https://www.covid19treatmentguidelines.nih.gov/therapies/antivirals-including-antibody-products/ritonavir-boosted-nirmatrelvir--paxlovid-/paxlovid-drug-drug-interactions
7. Miettinen OS. Proportion of disease caused or prevented by a given exposure, trait or intervention. *Am J Epidemiol*. 1974;99(5):325-332. doi:10.1093/oxfordjournals.aje.a121617
8. Khosravi A, Nazemipour M, Shinozaki T, Mansournia MA. Population attributable fraction in textbooks: time to revise. *Global Epidemiol*. 2021;3:100062. doi:10.1016/j.gloepi.2021.100062
9. Mansournia MA, Altman DG. Population attributable fraction. *BMJ*. 2018; 360:k757. doi:10.1136/bmj.k757
10. Carpenter J, Bithell J. Bootstrap confidence intervals: when, which, what? A practical guide for medical statisticians. *Stat Med*. 2000;19(9):1141-1164. doi:10.1002/(sici)1097-0258(20000515)19:9<1141::aid-sim479>3.0.co;2-f

#### eTable 1. Description of Electronic Health Record Data Sources.

| Health system | HealthPartners[a] | | Renown Health |
|---|---|---|---|
| Protocol name | myGenetics | ViEW Network™ | Healthy Nevada Project |
| Protocol number | HRN 001 | 0006-001 | 7701703417 |
| Geographic region | Minnesota and western Wisconsin | Minnesota and western Wisconsin | Northern Nevada |
| Total number of adults with SARS-CoV-2 infection | 1,301 | 1,776 | 928 |
| Number of patients meeting inclusion criteria | 1,089 | 1,224 | 724 |
| Date range of patients included | April 1, 2022–February 27, 2023 | February 28–June 23, 2023 | April 7, 2022–March 7, 2023 |
| Informed consent | Written informed consent obtained | Waiver of consent obtained | Written informed consent obtained |
| Data privacy | Limited dataset stripped of direct identifiers | De-identified dataset under expert determination | Limited dataset stripped of direct identifiers |

[a] HealthPartners patients were included across two different protocols (myGenetics and the ViEW Network™). To avoid possible overlap of the same SARS-CoV-2 infections among patients in both protocols, only myGenetics participants with infections prior to the first infection in the ViEW Network™ were included (before February 28, 2023). Patients could not be linked across protocols.

#### eTable 2. Definitions of High-Risk Conditions.

| Condition[a] | ICD-10-CM codes |
|---|---|
| Asthma | J45 |
| Cardiovascular disease | I01, I02, I05-I09, I21-I25, I27, I28, I31, I34-I37, I41-I44, I48, I50-I52, I71-I75, I79, I97.0, I97.1, M31.0, M31.1, M31.4, M31.6-M31.9, Q20-Q26, Q27.0, Q27.3, Q27.4, Q27.8, Q27.9, Q28, Z95, Z98.61 |
| Cerebrovascular disease | G45, G46, I60-I63, I68, I69 |
| Chronic kidney disease | I12, I13, N18, Z49, Z91.15, Z99.2 |
| Chronic lung disease | A15, A31.0, B39.1, B40.1, B41, B44.0, B44.1, B44.81, B45.0, B46.0, D86.0, E84, E88.01, I26, J40-J44, J47, J60-J66, J67.0-J67.8, J68, J70.1, J70.3, J81.1, J82.81, J84.0, J84.10, J84.111-J84.113, J84.115-J84.117, J84.17, J84.2, J84.8, J84.9, J96.1, J96.2, P27 |
| Dementia | F01-F03, G30 |
| Developmental or intellectual disability[b] | F71-F73, F78-F82, F84, F88-F90, Q00-Q07, Q76-Q79, Q85, Q87.4, Q90-Q93, Q96 |
| Diabetes | E10, E11 |
| Hematologic malignancy[c] | C81-C86, C88, C90-C96, D46 |
| HIV[c] | B20, B97.35, O98.7 |
| Liver disease | B18, I81, I85, K70-K77 |
| Mental health condition | F20-F25, F28-F34, F39 |
| Obesity[d] | E66.0-E66.2, E66.8, E66.9 |
| Other intrinsic immune disorder[c] | D70, D71, D72.0, D72.81, D72.89, D72.9, D80, D81, D82-D84, D89.1, D89.3, D89.4, D89.8, D89.9 |
| Solid malignancy | C00-C26, C30-C34, C37-C41, C43, C44, C4A, C45-C58, C60-C75, C7A, C7B, C76-C80 |
| Solid organ or stem cell transplantation[c] | D47.Z1, T86.0-F86.5, T86.81, T86.85, Z48.2, Z94.0-Z94.4, Z94.8, Z94.9, Z98.85 |
| Smoking (current/former)[e] | F17.20, F17.21, O99.33, Z72.0, Z87.891 |
| Rheumatologic or inflammatory disorder[c] | D86, E85, G35, G36, G37.1, G37.3, G37.8, G37.9, G61.0, G61.9, I40, J67.9, J84.01, J84.02, J84.09, K50-K52, L40.54, L40.59, L93.0, L93.2, L94.0, L94.2, M04-M08, M11-M14, M30, M31.3, M31.5, M32-M34, M35.0, M35.3, M35.8, M35.9, M46, T78.40 |
| **Immunosuppressive medication class[c]** | **Generic drug names** |
| Systemic corticosteroids | beclomethasone, betamethasone, budesonide, cortisone, deflazacort, dexamethasone, hydrocortisone, methylprednisolone, prednisolone, prednisone, triamcinolone |
| Immunomodulators | abatacept, acalabrutinib, adalimumab, alefacept, alemtuzumab, anakinra, azathioprine, baricitinib, basiliximab, belatacept, belimumab, belumosudil, blinatumomab, brentuximab vedotin, certolizumab pegol, certolizumab pegol, cyclosporine, daclizumab, daratumumab, eculizumab, epcoritamab, etanercept, everolimus, gemtuzumab ozogamicin, golimumab, ibritumomab tiuxetan, ibrutinib, infliximab, isatuximab, lenalidomide, lymphocyte immune globulin or anti-thymocyte globulin, methotrexate, muromonab-CD3, mycophenolate mofetil, mycophenolic acid, natalizumab, obinutuzumab, ocrelizumab, ofatumumab, pirtobrutinib, rituximab, rozanolixizumab, ruxolitinib, sirolimus, tacrolimus, tafasitamab, thalidomide, tocilizumab, tofacitinib, ublituximab, upadacitinib, ustekinumab, voclosporin, zanubrutinib |
| Chemotherapeutics | adagrasib, altretamine, amivantamab, amsacrine, arsenic trioxide, asparaginase, axicabtagene ciloleucel, azacitidine, belantamab, belantamab mafodotin, belinostat, belzutifan, bendamustine, bexarotene, |

| | |
|---|---|
| | bleomycin, brexucabtagene autoleucel, busulfan, cabazitaxel, calaspargase pegol, capecitabine, carboplatin, carmustine, chlorambucil, ciltacabtagene autoleucel, cisplatin, cladribine, clofarabine, cyclophosphamide, cytarabine, dacarbazine, dactinomycin, daunorubicin, decitabine, docetaxel, doxorubicin, enfortumab vedotin, epirubicin, eribulin, estramustine, etoposide, floxuridine, fludarabine, fluorouracil, gemcitabine, idarubicin, idecabtagene vicleucel, ifosfamide, inotuzumab ozogamicin, irinotecan, ivosidenib, ixabepilone, lisocabtagene maraleucel, lomustine, loncastuximab tesirine, lurbinectedin, mechlorethamine, melphalan, melphalan flufenamide, mercaptopurine, mirvetuximab soravtansine, mitomycin, mitotane, mitoxantrone, naxitamab, nelarabine, omacetaxine, omacetaxine mepesuccinate, oxaliplatin, paclitaxel, panobinostat, pegaspargase, pemetrexed, pentostatin, plicamycin, polatuzumab vedotin, pomalidomide, pralatrexate, procarbazine, romidepsin, sacituzumab, selinexor, streptozocin, tagraxofusp, talimogene laherparepvec, tazemetostat, teclistamab, temozolomide, teniposide, thioguanine, thiotepa, tipiracil/trifluridine, tisagenlecleucel, tisotumab, topotecan, trabectedin, tretinoin, uracil mustard, valrubicin, venetoclax, vinblastine, vincristine, vinorelbine, vorinostat |

Abbreviations: ICD-10-CM, *International Classification of Diseases, Tenth Revision, Clinical Modification*.

[a] High-risk conditions were defined by the presence of ICD-10-CM codes within the prior two years for asthma, cardiovascular disease, cerebrovascular disease, chronic kidney disease, chronic lung disease, dementia, developmental or intellectual disability, diabetes, hematologic malignancy, HIV, liver disease, mental health condition, obesity, other intrinsic immune disorder, solid malignancy, solid organ or stem cell transplantation, smoking (current or former), or by the presence of a drug prescription/administration record for immunosuppressive therapy within the prior six months (see details for immunocompromised definition below). All sub-codes of the listed ICD-10-CM codes were included.

[b] No patient had an ICD-10-CM code for a developmental or intellectual disability.

[c] Immunocompromised patients were defined as those with ICD-10-CM codes within the prior two years for hematologic malignancy, HIV, other intrinsic immune disorder, or solid organ or stem cell transplantation, or with a prescription/administration of immunosuppressive therapy in the prior six months. Systemic corticosteroids (oral or injectable) were only considered if an ICD-10-CM code for a rheumatologic or inflammatory disorder was present or if the drug was taken for ≥14 days (based on either start and stop dates or consecutive start dates, as accurate stop dates were not reliably captured in the electronic health record). All immunomodulators and chemotherapeutics were included. An ICD-10-CM code for a rheumatologic or inflammatory disorder alone was not sufficient for being defined as high-risk or immunocompromised.

[d] Obesity was also defined as body mass index ≥30 kg/m$^2$ using the most recent weight and height within two years.

[e] Although not an underlying condition, smoking (current or former) was used to define high-risk. In addition to ICD-10-CM codes, smoking was ascertained using social history documented in the electronic health record.

#### eTable 3. Definitions of Nirmatrelvir/Ritonavir Contraindications.

| Potential Moderate or Severe Drug-Drug Interactions[a] | |
|---|---|
| **Recommendation** | **Generic drug names** |
| Prescribe alternative COVID-19 therapy | amiodarone, bosentan, carbamazepine, clopidogrel, clozapine, dihydroergotamine, disopyramide, dofetilide, dronedarone, eplerenone, ergotamine, flecainide, glecaprevir/pibrentasvir, ivabradine, lurasidone, lumacaftor/ivacaftor, methylergonovine, midazolam (oral), phenobarbital, phenytoin, pimozide, primidone, propafenone, quinidine, rifampin, rifapentine, sildenafil, St. John's wort, tadalafil, tolvaptan, vardenafil, voclosporin |
| Temporarily withhold concomitant medication, if clinically appropriate | alfuzosin, aliskiren, atorvastatin, avanafil, colchicine, eletriptan, erythromycin, everolimus, finerenone, flibanserin, lomitapide, lovastatin, naloxegol, ranolazine, rimegepant, rivaroxaban, rosuvastatin, salmeterol, silodosin, simvastatin, suvorexant, tacrolimus, ticagrelor, tolvaptan, triazolam, ubrogepant, vorapaxar |
| **Condition[b]** | **ICD-CM-10 codes** |
| Severe renal impairment | I12.0, I13.11, I13.2, N18.4, N18.5, N18.6, Z49 |
| Severe hepatic impairment | K70.3, K70.4, K71.7, K72, K74.3, K74.4, K74.5, K74.6 |

Abbreviations: ICD-10-CM, *International Classification of Diseases, Tenth Revision, Clinical Modification*.
[a] Potential moderate or severe drug-drug interactions with nirmatrelvir/ritonavir were defined as the presence of one or more of the listed drugs within 30 days before through 7 days after the specimen collection date. Use of these drugs warrant either the prescribing of alternative COVID-19 therapy or the temporary withholding of the concomitant medication.
[b] Severe renal impairment and severe hepatic impairment were defined by the presence of ICD-10-CM codes within the prior two years. All sub-codes of the listed ICD-10-CM codes were included.
